# Hybrid diffuse optical appraisal of peripheral and cerebral changes in critically ill patients receiving red blood cell transfusion

**DOI:** 10.1101/2024.02.13.24302577

**Authors:** Susanna Tagliabue, Anna Rey-Perez, Lourdes Esposito, Andrés F. Jimenez, Sara Valles Angulo, Federica Maruccia, Jonas B. Fischer, Michal Kacprzak, Maria A. Poca, Turgut Durduran

## Abstract

**Background:** Red blood cells transfusions (RBCT) are utilized to restore normal values of hemoglobin concentration and hematocrit percentage in anemic patients. As expected, RBCT often leads to local and global alteration of blood flow (BF) and blood/tissue oxygenation which could have local deleterious consequences. This complicates its use and its dosage and there is no consensus on liberal versus restrictive RBCT in critically ill patients. Blood gas sampling is utilized to bring objectivity to RBCT which is a reliable systemic measure. However, it is also hypothesized that the knowledge about the dynamic response of selected organs could improve RBCT outcomes. We carried out a study using non-invasive hybrid diffuse optics (DO) to assess the RBCT effect on the brain and a peripheral muscle by evaluating microvascular BF, oxygen extraction fraction (OEF) and microvascular oxy-, deoxy- and total hemoglobin concentrations ([HbO_2_], [Hhb], [HbT]) in critically ill patients. We explored the DO’s ability to identify RBCT-induced significant alterations and to provide a quantitative description.

**Methods:** Critically ill anemic patients undergoing RBCT were recruited and monitored by hybrid DO. Blood gas samples were extracted to obtain arterial total hemoglobin concentration (Hgb) and hematocrit value. Optical signals, such as BF, OEF, metabolic rate of oxygen extraction (MRO_2_), [HbO_2_], [Hhb] and [HbT] were simultaneously measured at the cerebral and the peripheral tissues. The changes in these variables were investigated characterizing the distributions of the cerebral and of the peripheral post-RBCT variables.

**Results:** Fourteen out of fifteen recruited subjects were included. After RBCT, Hgb and hematocrit significantly increased (p<0.001). OEF significantly decreased both at peripheral and cerebral level (p<0.001, p<0.001). A significant increase was found in MRO_2_ (p=0.03, p<0.001), [HbT] (p=0.01, p<0.0001) and [HbO_2_] (p=0.008, p<0.0001) at both levels. BF significantly decreased only at the peripheral level (p<0.001). No change was encountered in [Hhb] (p>0.05). No statistical difference was found between cerebral and peripheral signals post-RBCT (p>0.05) apart from MRO_2_ (p=0.03, higher at peripheral tissue).

**Conclusions:** Hybrid DO detected tissue oxygenation improvement after RBCT, enabling a thorough examination. The potential for DO to quantify and alert changes of concern deserves further investigation.

## Background

Anemia is a common complication observed in critically ill patients admitted to the intensive care unit (ICU). In the ICU setting, anemia can result from severe bleeding after trauma, acute blood loss or hemorrhage during surgery and in non-bleeding but acutely ill patients experiencing a fast decrease in the hemoglobin concentration in blood [1]. Typically, anemia is defined by low arterial oxy-hemoglobin concentration (HgbO_2_) in blood or low hematocrit (HCT) percentage (about 8-10 g/dL [2, 3] and 30% [4] respectively) relative to normal values (about 14-15 g/dL and 35-50% respectively)^[1]^. This is quite common for the general ICU critically ill population within the first three days after admission.

In anemic condition, low HgbO_2_ translates into reduced oxygen supply, release and diffusion into tissues. This affects the blood flow (BF, cerebral blood flow (CBF) when related to the brain, peripheral blood flow (PBF) when related to peripheral parts of the body) [2], due to its relationship with the oxygen delivery (DO_2_), which is the product of BF and arterial oxygen content (CaO_2_) [4, 5]. As such, anemia can have deleterious effects on the patient health [2, 5] and extended anemia must be avoided [6] with interventions to restore its normal levels as soon as possible.

As an intervention, patients are generally transfused with units of red blood cells (RBC) [1] and often benefit from this RBC transfusion (RBCT). This intervention is accepted by most clinicians [1, 4], with the objective to restore the content of HgbO_2_. For the purposes of this manuscript, among other things, RBCT alters the BF [1, 6] and other physiological parameters directly related to them, such as the arterial total hemoglobin concentration (Hgb), the tissue oxygen saturation (StO_2_), the metabolic rate of oxygen (MRO_2_) and the oxygen extraction fraction (OEF). In the case of healthy brain, cerebral autoregulation and other protective mechanisms ensure that, at a local level, these changes are not impairing the brain function. For the non-critical organs, such as peripheral muscles, this may not be the case and large, possibly unsustainable changes may be observed. In other words, the balance of local hemodynamics and metabolism can be altered by RBCT.

Nonetheless, the indications for RBCT for critical care patients are subject of considerable debate among clinicians. For example, the “Transfusion Requirements in Critical Care (TRICC)” study, published in 1999 [5, 7] and reviewed in 2012 [8], questioned whether transfusions are appropriate, especially in ICU patients with acute brain injuries, and their risk-benefit ratio [2, 4–6, 9].

Several studies have reported side effects of RBCT in traumatic brain injury (TBI) patients [4] and recent guidelines recommend a more restrictive transfusion strategy for them, with a HgbO_2_ threshold of 7g/dL or lower [1–4, 8, 10, 11]. In fact, although in TBI patients anemia is a recognized contributing factor to secondary injury [4, 12] and is associated with increased morbidity, poorer neurological outcome [2, 4, 6] and higher mortality [6], their brain is more vulnerable to the hypoxic damage that may be caused by RBCT [2, 4, 12, 13]. This may be because cerebral autoregulation [2, 4], the systemic cardiovascular response and neuroprotection mechanisms are frequently impaired [2].

Furthermore, TBI patients usually present low CBF [2, 4, 6, 14], which increases the risk of ischemia whenever the oxygen supply cannot meet anymore the demand [2]. This may be worsened by RBCT if blood is directed from the brain to the peripheral organs in response to the treatment. The identification of the hemodynamic and metabolic evolution that RBCT induces in the brain versus peripheral muscle could indicate to which extent the vulnerable brain of neurocritical patients is affected and with respect to the peripheral system.

For this reason, advanced photonics methods based on non-invasive hybrid diffuse optics (DO) that utilize near-infrared light can be exploited. Hybrid DO, comprising time-resolved spectroscopy (TRS) and diffuse correlation spectroscopy (DCS), could provide more complete overview of the physiological alterations. DCS offers insight about the BF (CBF and PBF), while TRS on the StO_2_, microvascular oxy- and deoxy-hemoglobin concentrations ([HbO_2_] and [Hhb]), total microvascular hemoglobin concentration ([HbT]), OEF (COEF when cerebral, PEOF when peripheral). Together they convey information about the MRO_2_ (CMRO_2_ when cerebral, PMRO_2_ when peripheral). The literature on the effects of RBCT on these signals is quite limited [15]. Continuous-wave near-infrared spectroscopy (NIRS) is the most commonly employed method to measure blood oxygen saturation [5, 15] with few studies on children (i.e. premature babies) [16–22] and fewer in adults [9, 23–26].

We have conducted a pilot study where both local cerebral and peripheral continuous monitoring were performed by hybrid DO throughout RBCT in critically ill patients. This study was exploratory and observational, seeking to inspect cerebral and peripheral hemodynamic and metabolic biomarkers and aiming at demonstrating the capability of DO spectroscopic techniques in quantifying changes due to RBCT.

### Study hypotheses

In this study, we aimed at describing and quantifying the response to RBCT for all optically-derived physiological signals. We have hypothesized that RBCT affects them and wanted to verify whether these changes were significant with respect to initial, baseline values prior to the RBCT. We did this both at peripheral and cerebral level, to further inspect whether the response and the amount of their response differed between sites. This allows for the assessment of potential deleterious effects after RBCT.

## Material and methods

### Subjects and clinical protocol of management

Critically ill patients aged 16-72 admitted to the neurotrauma ICU of the Vall d’Hebron University Hospital (VHUH) between June 2019 and July 2021 that required blood transfusion were included in the study. Despite being created to admit trauma patients, this ICU actually sometimes admits critical patients of different etiologies, such as patients with cerebral hematomas, cerebrovascular damage and large burns, either with a trauma or not, that often lose fluids and blood due to their condition. Among all critically ill patients, we have recruited: 1) polytrauma patients with (abnormal admission computed tomography (CT) scan) or without (normal CT scan) moderate or severe TBI (Glasgow coma scale (GCS) ≤ 13), 2) patients with ischemic or hemorrhagic stroke and 3) patients with severe burns.

Severe TBI patients were treated following the Brain Trauma Foundation (BTF) guidelines [27] and the neuromonitoring protocol included continuous intracranial pressure (ICP) and cerebral oxygenation (PbrO_2_) implanted in the most injured hemisphere. The rest of the patients were treated under standard care of critically ill patients and the above TBI guidelines were transposed to them when necessary. Anemia was defined as a HgbO_2_ ≤ 13 g/dL for adult males and ≤ 12 g/dL for adult non-pregnant females. Below these oxy-hemoglobin values, a blood transfusion was indicated. RBC bags were provided for allogenic transfusion by the hospital transfusion services (blood bank), tested for compatibility, verified for blood type and cross-match, that were completed within the prior 96 hours. Whole blood (RBC, platelets, fluids, etc.) was administered after a health care provider request.

### Study protocol

A schematic of the measurement protocol and probe positioning is provided in **Figure 1**. One optical probe was placed on the forehead of the recruited subject, while at bedside (cerebral placement). CT scans were checked prior to the measurements whenever available to inspect the tissue and decide the optimal positioning. Accordingly, the hemisphere more easily accessible and/or without underlying trauma was always preferred for the probe placement. A second probe was preferably placed on the *quadriceps femoris* or, if not accessible, on the *brachioradialis* muscle (peripheral placement).

**Figure 1:**
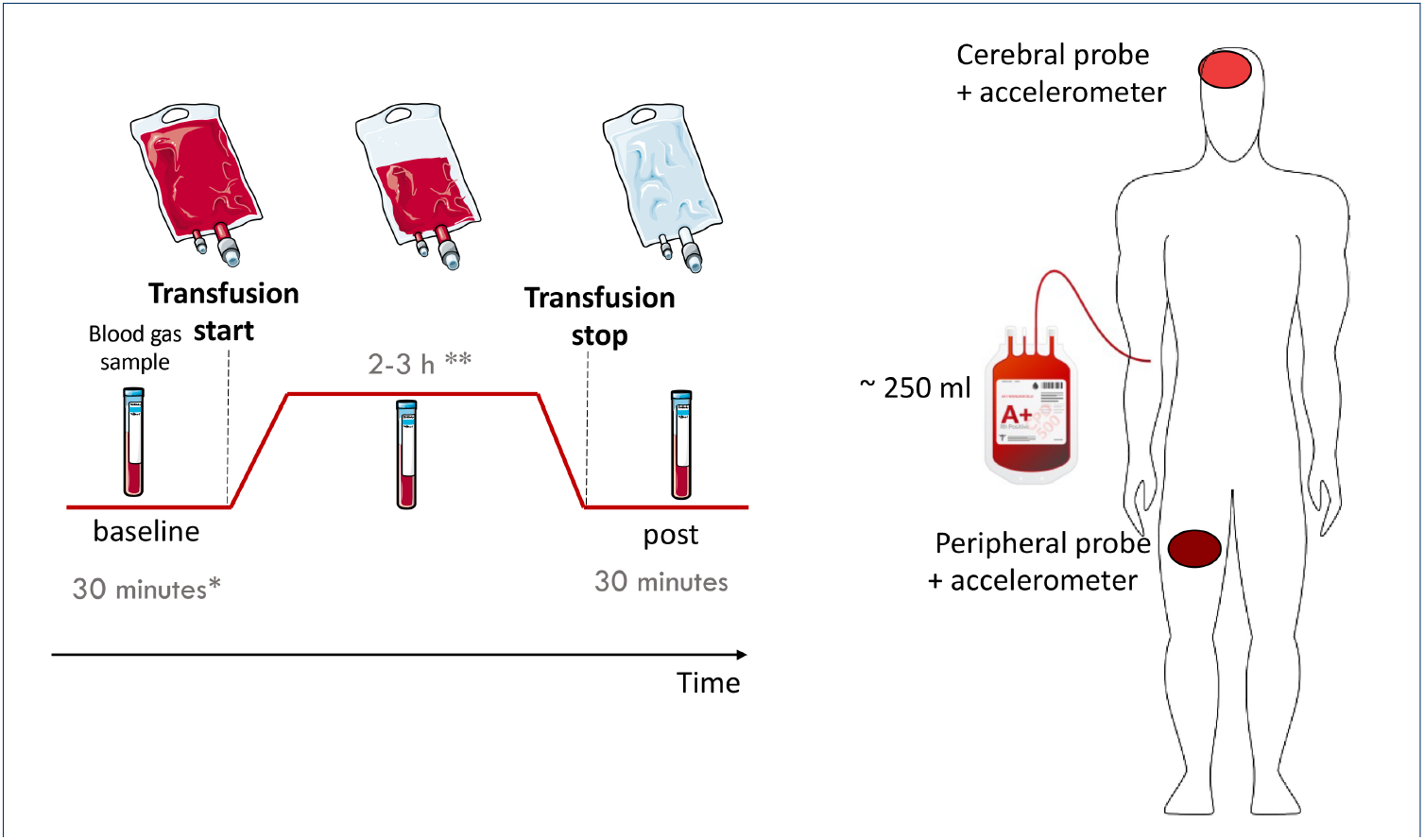
Study protocol: following a baseline pre-RBCT period (* generally about 30 minutes, but adjusted to the patient’s needs), the RBC transfusion started (** 2-3 hours duration according to the bag volume and/or transfusion rate decided by the nurse) and was measured until finished, when additional 30 minutes post-transfusion were monitored. Three blood gas samples were taken during each period of the measurement. As depicted on the right, one probe was placed on the forehead of the subject, while the second at peripheral level, generally on the *quadriceps femoris*. This Figure was created by modifying and merging images from Servier Medical Art (http://smart.servier.com/), part of Laboratoires Servier, licensed under a Creative Commons Attribution 3.0 Unported License (https://creativecommons.org/licenses/by/3.0/).

After the positioning of the optical probes, critical care personnel re-verified the blood bag compatibility with the patient and the standard transfusion procedure was followed.

The continuous monitoring of the patients was carried out before, during, and after transfusion with a protocol of 30 minutes of baseline (pre-RBCT), 2-3 hours-long RBCT and 30 minutes of observational post-transfusion monitoring (post-RBCT). The timing of each period was adjusted according to the situation as illustrated in **Figure 1**.

Standard vials of blood samples were also extracted from an artery (generally the *arteria radialis*) through a catheter during the protocol at baseline, when the half the blood bag was transfused and during the observational final period, for a total of three times. In the ideal scenario, the initial and final blood samples were taken about 10 minutes after each protocol part begun, but was adjusted according to the decisions of the medical personnel.

In order to standardize the protocol and its duration, we have decided to carry out our study only after one single RBCT even in those cases where more blood bags were indicated to restore the patient hemoglobin values.

### Clinical monitoring

Standard bedside monitors were used to collect physiological and clinical parameters. Mean arterial blood pressure and heart rate were measured by available sensors and were recorded by the critical care monitor (IntelliVue MX800 or MX750, Koninklijke Philips N.V., Netherlands). Peripheral arterial oxygen saturation was measured by a standard pulse-oximeter (Nellcor DS100A-1, Medtronic, United States of America) through a capnograph with a pulse-oximeter unit (Capnostream^*T M*^ 20p, Medtronic, United States of America).

A dedicated software (LabChart, version 6 and 8.1, ADInstruments Ltd, United Kingdom) and hardware (PowerLab, ADInstruments Ltd, United Kingdom) recorded all digitized vital signals at a sampling frequency of 400 Hz alongside a synchronization signal from the hybrid DO device.

The samples of arterial blood were analyzed by a co-oximeter (GEM Premier 4000, Werfen, Spain) to obtain the levels of arterial carbon-dioxide (PaCO_2_), HgbO_2_, arterial reduced hemoglobin concentration (Hgbr), arterial total hemoglobin concentration (Hgb) and HCT.

Patient demographic information was also collected, including sex, age and etiology.

### Optical monitoring

A hybrid DO device, previously described in References [28, 29] and similar to Reference [30], comprising of diffuse correlation spectroscopy (DCS) and time-resolved spectroscopy (TRS), was used for the non-invasive optical measurements. Both techniques are based on diffuse near-infrared spectroscopy methods [31]. DCS quantifies the motion of scatterers inside of the probed tissue volume which measures BF [32]. TRS recovers the wavelength dependent absorption and scattering of the probe tissue volume [31, 33]. In human tissues, these quantities allow one to retrieve [HbO_2_], [Hhb], [HbT] and *StO*_2_ [31, 33]. Further details about the optical device, probe adhesion and data acquisition can be found in Reference [28]. Real-time movement monitoring was employed using accelerometers on programmable boards placed at the two distinct patient placements, above the probes.

Events were marked within the software routine and data were synchronized with high precision with the ones provided by the clinical monitors as described above.

### Data evaluation

DO analysis for the collected data followed the standard procedures for fitting methods for both TRS and DCS techniques [33–35] and making use of Matlab (Release 2018b, The MathWorks, Inc. Natick, Unites States of America). The optically-derived parameters of interest in this study were: [HbO_2_], [Hhb], [HbT], BF, OEF, MRO_2_. Again, each acronym is preceded by a ‘C’ when referred to the cerebral tissue and by a ‘P’ when peripheral tissue. Probe placement was described above and shown in **Figure 1**.

Strict rejection criteria were applied to the data according to the signal-to-noise ratio (SNR) and the measurement session quality as detailed in Reference [28]. Additionally, accelerometers information was inspected when an artifact was suspected from the marks. In this work, the SNR for TRS was defined as the ratio between the maximum of the TRS distribution of time-of-flight, which is the retrieved curve built by the photons distribution after the initial laser pulse traverses the tissue, and the standard deviation of a background portion of the same signal prior to the curve. The threshold value was SNR>10. DCS measurements were rejected if the averaged count-rate of the detector channels to build the autocorrelation function was lower than 10 kHz and *β* parameter was lower than 0.4 [34].

The non-rejected time-traces of the mentioned variables were then synchronized with the clinical signals by exploiting the common time-base signal. Further parameters, like OEF and MRO_2_, based on both optical and clinical data, were derived as described in References [28, 34, 35].

Since recent studies showed the need to apply corrections to DO signals calculations when HCT changes significantly during measurements [36, 37], a correction in the MRO_2_ time-traces calculation was implemented. Accordingly, MRO_2_ calculation was also corrected by the HCT value measured by the blood gas analysis and similarly to Reference [38]. To do so, in the following expression:

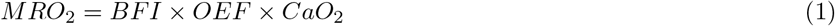

the arterial oxygen content (CaO_2_) was computed as

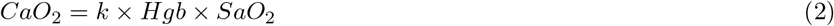

where *k* was assumed equal to 1.36 mLO_2_/1gHgb for mammalians and the arterial total hemoglobin concentration was expressed as

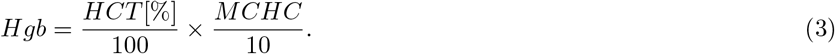

In the latter equation, MCHC is the mean corpuscular hemoglobin concentration and was assumed to be the averaged value 340 g/L. For SaO_2_, the time-trace collected by the additional monitors was used. Since in this study HCT was available in three moments only, the computation of MRO_2_ was done separately for each period of the protocol with its respective HCT value kept as a constant.

All the optically derived hemodynamic signals were calculated for cerebral and peripheral areas below the probe placements separately. PBF, POEF, PMRO_2_ and their cerebral counterpart time-traces were referred to their pre-RBCT baseline period by dividing the entire time-trace by the pre-RBCT mean value. The baseline pre-RBCT lasted from the beginning of the recording to the beginning of the RBCT, marked as “transfusion start”. In this case, an “r” was added to describe these variables as relative (i.e. relative CBF = rCBF). Instead, [HbT], [HbO_2_] and [Hhb] were calculated as difference changes and, therefore, each time-trace was subtracted by its average over the baseline pre-RBCT period. This led to Δ[HbT], Δ[Hhb] and Δ[HbO_2_] for both peripheral and cerebral probes.

The data related to vital signals collected by external available monitors were read in software during post-processing and synchronized with the optical time-traces. All data were down-sampled to 10 second bins.

Marked events were reported on all physiological time-traces. This led to further artifact identification and removal corresponding to periods of patient manipulation due to clinical procedures. Similarly, the visual inspection of data allowed the identification of features such as movement artifacts.

Relevant blood-derived parameters obtained by the output of the blood gas analyzer were collected for each subject in a database together with all demographic and clinical patient information.

### Statistical analysis

All statistical analyses were carried out with R (v4.0.1, R Core Team, USA) and the integrated development environment RStudio (v1.1.5042, RStudio, Inc., USA). In particular, the *nlme* v. 3.1-152, *lme4* v. 1.1-27, *lmerTest* v. 3.1-3 and *stats* v. 3.6.3 packages were utilized.

Descriptive statistics was computed for each variable. Clinical variables and demographic information (i.e. sex) were summarized as mean(standard deviation (SD)) or proportions (i.e. males:females as M:F).

Variables obtained at specific time-points by blood gas samples (up to three times per measurement session), i.e. Hgb, HCT and PaCO_2_, were summarized by mean(SD) both before (pre-) and after (post-) transfusion. Pre-transfusion values were compared to post-transfusion ones by Wilcoxon paired tests.

Continuous normally distributed optically-derived hemodynamic time-traces underwent a windowing procedure in order to define pre- and post-transfusion values, both at the cerebral and peripheral levels. The pre-transfusion window was selected lasting the entire pre-RBCT period, prior to marking the beginning of the RBCT. The post-transfusion window included the six minutes after the marked event of RBCT end. The values within the windows were averaged and collected separately into two distributions, one for the cerebral and one for the peripheral placement.

For each variable, the difference between the post-transfusion distribution of mean values with respect to its pre-RBCT distribution was tested by unpaired Wilcoxon test: the mean of the pre-RBCT was subtracted to the post-RBCT distribution (providing a relative mean) and its difference from zero tested. For the distributions of variables which were expected to be increased after transfusion, one-sided Wilcoxon tests were used to check whether the relative change was greater than zero (with “greater” option for the test applied), while one-sided test to check whether the relative mean was less than zero (with “less” option employed) for those variables expected to show a decrease. Significance was attributed when p-values (p) were lower than 0.05 and all values lower than 0.001 were reported only as p<0.001. Furthermore, the paired Wilcoxon test was used to compare cerebral and peripheral pre-transfusion distributions of values to make sure that there were no initial differences (thus discarding cases where one placement was not available, i.e. cerebral missing but peripheral available). Paired Wilcoxon tests were also used to test the difference between cerebral and peripheral data distributions post-transfusion. Boxplots (median, first and third interquartile ranges (IQR)) for all pre- and post-RBCT distributions of mean values and variables, separated between cerebral and peripheral placements, were depicted to visualize the data. The median values with respect to the baseline pre-RBCT were used to identify the direction of the change for the distribution, for instance an increase if it was greater than the baseline and statistically significant.

## Results

### Population characteristics

Fifteen subjects were recruited (see **Table 1**) with ten patients with polytrauma, three with ischemic or hemorrhagic stroke and two with severe burns. The data from one subject had to be discarded due to poor SNR in the optical measurement (according to the criteria described above) both on the head (effect of the trauma) and the peripheral muscle (scar tissue due to an old burnt). Furthermore, two data sets from the probe in the cerebral placement were discarded due to poor SNR.

**Table 1:**
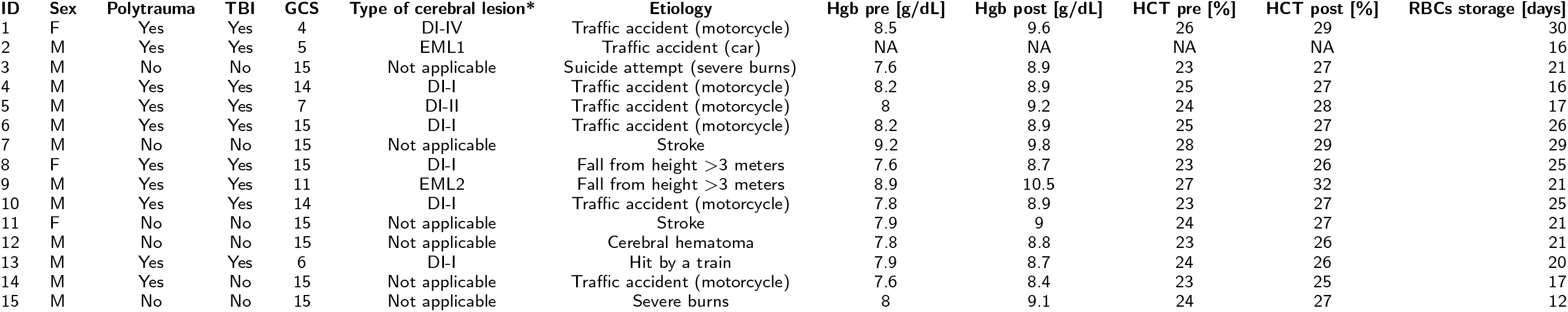
Demographic and clinical data (n=15). F: Female; GCS: Glasgow Coma Scale score, value at hospital admission; Hgb pre: blood hemoglobin concentration before transfusion (g/dL); Hgb post: blood hemoglobin concentration after transfusion (g/dL); M: Male; RBCs: Red blood cells; TBI: Traumatic brain injury; * Type of cerebral lesion according to the Traumatic Data bank Classification [57] in the computed tomography scan closest to the study. Diffuse injury I (DI-I): no visible intracranial pathology seen on CT scan; Diffuse injury II (DI-II): cisterns are present with midline shift 0-5 mm and/or high or mixed-density lesions ≤ 25cc: Diffuse injury III (DI-III): cisterns compressed or absent with midline shift 0-5 mm and/or high or mixed-density lesions ≤ 25cc; Diffuse injury IV (DI-IV): midline shift >5mm without high or mixed-density lesion > 25cc; Evacuated mass lesion (EML): any lesion surgically evacuated; Non evacuated mass lesion (NEML): high or mixed-density lesion > 25cc not surgically evacuated. 1: Epidural hematoma. 2: Subdural hematoma. Patient ID = 3 was discarded. NA: not available.

Consequently, fourteen subjects were included in the following analysis out of which all the measurement sessions had peripheral measurements of the optical variables (time-traces), while twelve also had cerebral hemodynamic time-traces. Moreover, in one subject the peripheral measurement had be to carried out on the *brachioradialis* muscle due to inaccessibility of the *quadriceps femoris* muscle.

In one subject, the vial of blood sample was extracted only prior to the measurement due to the care provider decision. In this case, for the hematocrit correction of MRO_2_, it was decided to use as post-transfusion value the average over all of the other subjects’ post-transfusion HCT values which had started from the same initial HCT value as this subject.

The mean age of the fourteen included patients was 39(19) years (median: 35, minimum: 16, maximum: 72 years) and the proportion of males to females was 11:3. **Table 1** summarizes the demographic and clinical data of the patients. Descriptors for the variables related to the blood gas samples are summarized in **Table 2**. For the included subjects, the average RBC bag volume was 306(22) mL and the average bag age was 21(5) days.

**Table 2:** Results in terms of mean(SD) for Hgb, PaCO_2_ and HCT values obtained by blood gas analysis pre- and post-transfusion (pre-RBCT and post-RBCT). Post-transfusion values were tested against pre-transfusion values to verify whether had undergone a statistically significant change (significant when the p-value (p) <0.05). Additionally, an arrow upwards indicates that the post- values increased (median value for the distribution higher than for the pre-transfusion values), while a down-wards arrow stays for a decrease. Hgb: Arterial total hemoglobin concentration, measured by blood gas analysis; PaCO_2_: Arterial pressure of carbon dioxide; HCT: Hematocrit; RBCT: red blood cell transfusion. Dir.: direction of change.

| Variable | Pre-RBCT | Post-RBCT | Dir. | p-value |
| --- | --- | --- | --- | --- |
| Hgb [g/dL] | 8.1(0.48) | 9.1(0.55) | ↑ | p<0.001 |
| PaCO<sub>2</sub> [mmHg] | 39.4(4.3) | 40(4.5) | - | p=0.22 |
| HCT [%] | 24.5(1.6) | 27.4(1.8) | ↑ | p<0.001 |

### Representative data during transfusion

A representative set of data (male, age range 18-23 y.o., with polytrauma and severe TBI) is presented in **Figure 2** (case 2 in **Table 1**) where darker curves show data from the peripheral muscle (*rPBF, rPOEF, rPMRO*_2_) and the lighter curves correspond to data from the brain (*rCBF, rCOEF, rCMRO*_2_). The start and the end of the RBCT are indicated by vertical dashed lines. Additional events, such as blood gas sampling and movements, are depicted by lighter vertical lines. Initial Hgb, HCT and PaCO_2_ values were 7.9 g/dL, 26 % and 36 mmHg respectively.

**Figure 2:**
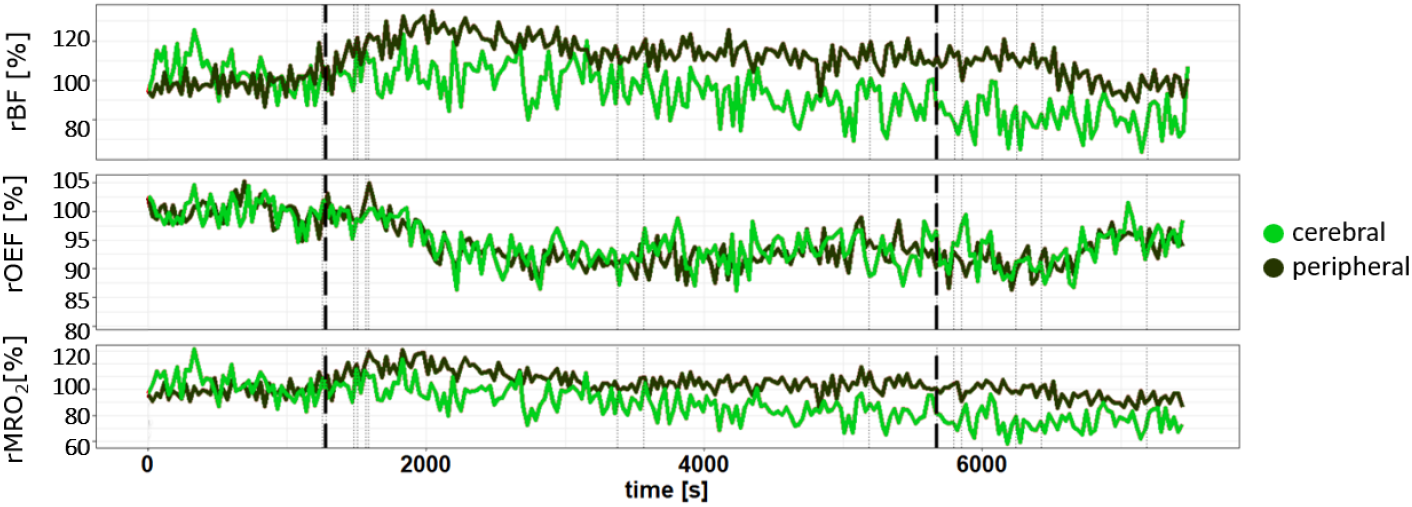
Time-traces collected during the RBCT of a patient for an entire protocol, comprising baseline pre-RBCT, blood unit transfusion and post-transfusion period. Non-invasive signals are depicted with peripheral and cerebral recordings color-coded as shown in the legend. The beginning and end of the process are highlighted by thick dashed vertical lines, synchronized in time for all variables. Additional events marked during the protocol are represented by vertical lines.

By eye, it can be seen that there is a general trend for the rCBF to decrease with respect to the pre-RBCT period, while the rPBF has an increasing trend. Instead, rOEF shows a decreasing trend for both peripheral and cerebral signals. The response of the metabolic rate of oxygen extraction is more subtle with a small decrease in rCMRO_2_ by the end of the protocol.

### Pre- and post-transfusion comparisons

As far as blood gas samples-related variables are concerned, the results of the comparison between pre-RBCT and post-RBCT distributions of mean values to check whether a significant change occurred are provided in **Table 2**, where the p-values and direction of changes are reported. RBCT caused a significant increase in the Hgb by about 0.9 g/dL (p<0.001) and HCT measured by blood gas samples by about 3% (p<0.001). PaCO_2_ did not show any significant change (p=0.22).

In relation to the tests for pre- and post-transfusion distributions from the hemodynamic time-traces, the results are detailed in **Table 3** and depicted in **Figure 3**. Differences between cerebral baseline pre-RBCT distributions and peripheral baseline pre-RBCT distributions for all variables were not statistically significant (p>0.05) and were omitted from the **Table**. All post-transfusion test results are provided together with the direction (Dir.) of the change with respect to the baseline. Moreover, the results of the tests between the post-RBCT cerebral values and the peripheral values are reported in the third column.

**Table 3:** Summary of the results of all tests carried out for the optically derived variables (rBF, rOEF, rMRO_2_, Δ[Hb*O*_2_], Δ[Hhb], Δ[HbT]). Under the column ‘Cerebral’, the post-transfusion values were tested for difference with respect to baseline. The results concerning the post-transfusion muscle-related values are reported below ‘Peripheral’, again tested with respect to baseline. Post-transfusion values for the cerebral level were tested for significant difference with respect to the post-transfusion values at peripheral level in the last column (‘Cerebral versus peripheral’). Significance was defined as p-values lower than 0.05. The direction (Dir.) of the change of the median value for the distribution is represented by an arrow: upwards if it increased, downwards if decreased. * in this case the arrow indicates that the peripheral level was significantly higher than the cerebral level after RBCT. rBF: relative blood flow; rOEF: relative oxygen extraction fraction; rMRO_2_: relative metabolic rate of oxygen; Δ[HbT]: Change in [HbT]; Δ[HbO_2_]: Change in [HbO_2_]; Δ[Hhb]: Change in [Hhb].

| Variable | Cerebral |  | Peripheral |  | Cerebral VS peripheral |  |
| --- | --- | --- | --- | --- | --- | --- |
|  | p-value | Dir. | p-value | Dir. | p-value | Dir. |
| rBF | 0.07 | - | <0.001 | ↑ | 0.1 | - |
| rOEF | <0.001 | ↓ | <0.001 | ↓ | 0.2 | - |
| rMRO<sub>2</sub> | 0.03 | ↑ | <0.001 | ↑ | 0.03 | ↑ * |
| Δ[HbO<sub>2</sub>] | 0.008 | ↑ | <0.001 | ↑ | 0.9 | - |
| Δ[Hhb] | 0.2 | - | 0.1 | - | 0.2 | - |
| Δ[HbT] | 0.01 | ↑ | <0.001 | ↑ | 0.5 | - |

**Figure 3:**
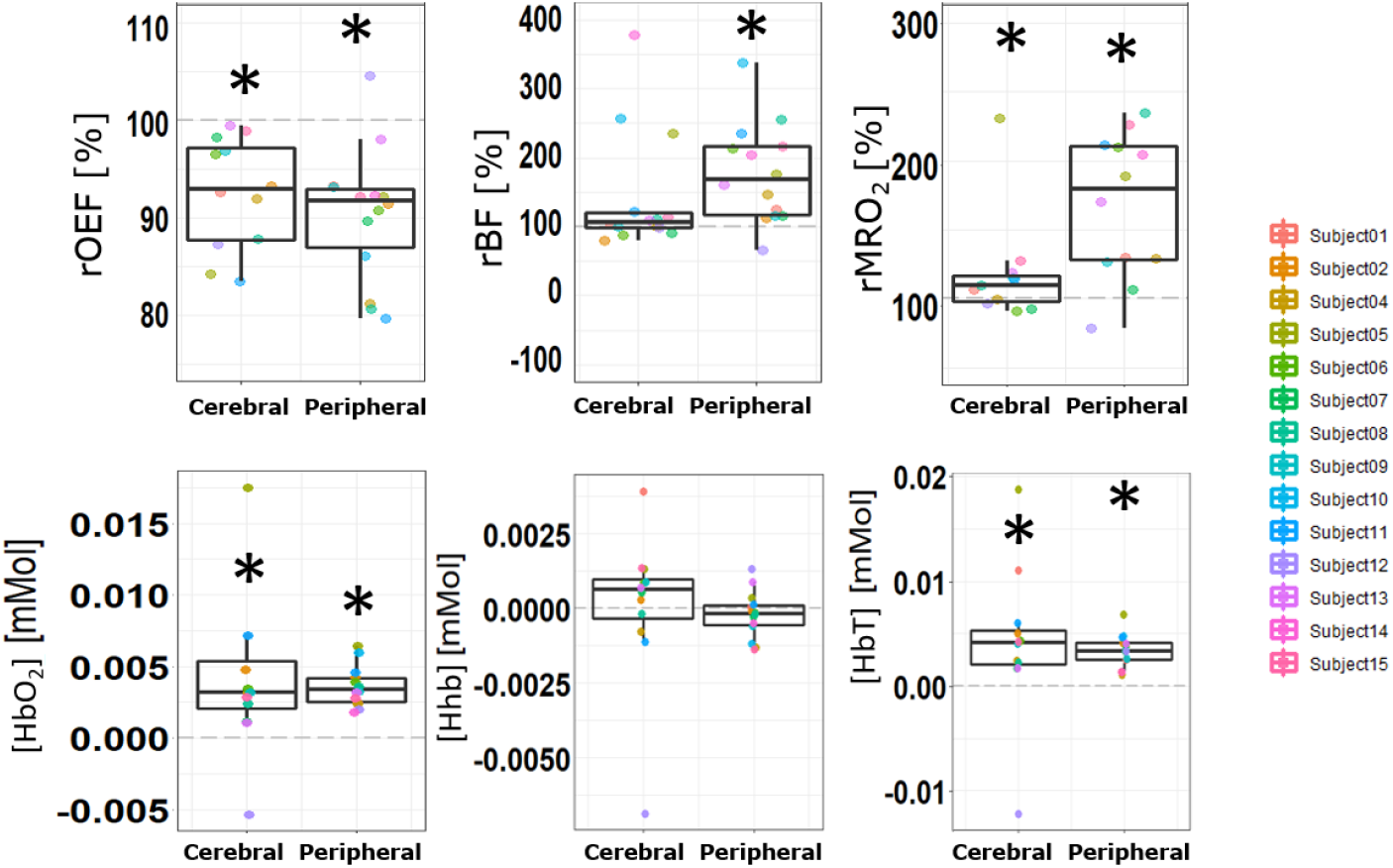
Graphs representing the post-transfusion data distribution for all optically-derived variables and divided into cerebral and peripheral muscle. For each distribution of mean values, its boxplot is represented (median, first and third quartiles). Dashed grey lines are drew indicating the baseline pre-RBCT values for the distributions, which they are relative to and compared with. Subjects are color coded in the same way for all variables. An asterisk on the top of a boxplot indicates that the distribution tested significantly different than the baseline. No significant difference was found between peripheral and cerebral post-transfusion values apart from the metabolic rate of oxygen.

RBCT provoked a significant increase (p<0.001) in rPBF by 68% (by comparing the percent change between the median post- to median pre-values), while no significant change in rCBF was observed (p=0.07). rPOEF median value fell significantly by 8% (p<0.001) and rCOEF by 7% (p<0.001) with respect to their pre-RBCT values. The median rPMRO_2_ significantly raised by 89% (p<0.001) and rCMRO_2_ by 10% (p=0.03), with a significantly greater median increase in the muscle by 79% than in the brain (p=0.03). No significant differences were found in the post-RBCT comparison for the other hemodynamic signals (p= 0.1 for rBF, p=0.2 for rOEF).

Both Δ[Hb*O*_2_] and Δ[HbT] had a significant increase both at cerebral and peripheral levels, with similar median shift with respect to baseline pre-RBCT values (as visible in **Figure 3**). Δ[Hb*O*_2_] increased by 0.0040(0.0053) mMol on the brain and 0.0036(0.0014) mMol on the muscle; Δ[HbT] raised by 0.0041(0.0071) mMol on the brain and 0.0034(0.0016) mMol on the muscle. Instead, Δ[Hhb] had no significant changes (p=0.2 and p=0.1 for cerebral and peripheral placements, respectively). No statistical difference was found between post-RBCT cerebral versus peripheral placement for any of these variables.

## Discussion

The study aimed at demonstrating the usefulness of hybrid DO to quantify the physiological changes after RBCT and in providing a preliminary characterization of the response at both the peripheral muscle and the brain in a small, pilot population of critically ill patients. We have obtained a rich array of signals. Since the RBCT response for the invasive and standard variables is well-documented, we focus on discussing the results of the optical monitoring with respect to the literature.

RBCT provoked a significant increase in rPBF, while no significant change in rCBF was observed. However, this is in contrast with the results obtained on the brain in ill but young populations (i.e. sickle cell disease, very preterm infants) where a decrease in the CBF was found after the intake of blood [15, 20, 39]. For instance, in Reference [20], the decrease of cerebral blood volume (CBV) was 0.5 mL/100g, while in Reference [39] CBV decreased by median 18.2% and CBF by median 21.2%. However, we acknowledge that the populations, the age, the origin of their anemia and the etiology are not fully relatable and may account for these discrepancies. As for alternative monitoring methods, for example, a reduction by 18% in the blood flow velocity measured by transcranial Doppler ultrasound (TCD) was found in stroke patients after 3h post-RBCT both on infarcted and non-infarcted regions, with significantly lower baseline values for the infarcted areas [40]. Nonetheless, in another study involving subarachnoid hemorrhage (SAH) patients monitored by ^15^O-positron emission tomography (PET), post-RBCT global CBF values remained unchanged [41], similar to our findings. Differences in the post-RBCT CBF response between children and adults with sickle cell anemia were also found by magnetic resonance imaging (MRI) and magnetic resonance angiography [42]: post-RBCT CBF did not significantly change in the adults (N=16), while it significantly decreased in the children (N=10, with -22.3 ml/100g/min in 9/10 participants). In Reference [43], CBF evaluated in children with sickle cell anemia by MRI with arterial spin labeling significantly decreased by 6.5 mL/100g. These findings are mostly in contrast with our results. However, a possible explanation may lie again in the etiology itself and the age of the participants. Moreover, the differences may be due to the fact that we have decided to monitor only the transfusion of a single RBC bag, the first one, even when more units had been assigned to the patient. This was done in order to be consistent among patients and be able to compare the amount of change of the physiological signals. According to literature, greater changes are expected when more bags are transfused [44].

Concerning TRS-related results for oxygenation and hemoglobin, we will try to discuss them altogether. Moreover, since several NIRS studies characterized the tissue oxygen saturation (StO_2_) rather than OEF, but which is generally inversely related to the OEF, we will use it as well as an inverse comparison to our findings. In our study, at cerebral level we encountered a significant decrease in rCOEF, a significant increase in Δ[Hb*O*_2_] and Δ[HbT], no change in Δ[Hhb]. For rCOEF, this is in agreement with literature and was mainly confirmed in young populations after RBCT [5, 15, 19, 45], although not always a significant change was found [26]. rCOEF significant decrease by 0.06% by CW-NIRS was reported, for instance. Studies using other modalities reported a significant decrease in post-RBCT rCOEF, which was observed both in children and adults with sickle cell anemia evaluated by MRI (−0.1)[42], in children with sickle cell anemia assessed by asymmetric spin echo sequence MRI technique (−2% for the whole brain) [43] and in SAH patients by PET (−0.11)[41]. These results are consistent with ours. Our results are also strengthened by several studies highlighting a significant increase in cerebral StO_2_ [17–21, 25, 46–48]. In Reference [17], cerebral regional StO_2_ measured by CW-NIRS increased by 17% in infants and by 4.2% in patients undergoing transfusion during aortic or spinal surgery [46]. Only in Reference [26], McCredie et al. did not detect any significant change in cerebral StO_2_ by bifrontal NIRS measurements in TBI after RBCT. Only one study used a TRS device in infants [20] and found the same: cerebral StO_2_ increased by about 2% after transfusion. As for Δ[Hb*O*_2_] and Δ[HbT], our findings are in line with other studies using NIRS [16, 20, 26, 47]. For instance, in Reference [16], Δ[Hb*O*_2_] was 0.026 mMol and Δ[HbT] was 0.02 mMol, and were in the same direction as our study, although with greater absolute value and measured by a frequency domain tissue oximeter.

In our study, at the peripheral level, we have found a significant decrease in rPOEF, a significant increase in Δ[Hb*O*_2_] and Δ[HbT] but no significant change in Δ[Hhb]. rPOEF result is in agreement with literature [5, 15, 19, 45], where a 0.13% significant decrease in the abdomen was found, for example, comparable to ours. Conversely, a lot of studies highlighted a significant increase in StO_2_ peripheral level [17–21, 25, 46–48], which is also in accordance with our study. For instance, peripheral StO_2_ increased by 17% in infants [17] and by 1.6% in patients undergoing transfusion during aortic or spinal surgery [46]. The peripheral increase in Δ[Hb*O*_2_] and Δ[HbT] was confirmed by NIRS literature [16, 20, 26, 47]. Regarding Δ[Hhb], similarly was found interscapularly by CW-NIRS [16]. However, Δ[Hhb] was often reported to decrease in literature.

It is possible to speculate that the variations in [Hhb] were too small and could have probably been more relevant in case of full restoration to normal haemoglobin concentration and hematocrit values, which were not reached by the end of the measurements due to our protocol choice. Furthermore, some studies highlighted RBCT failure in raising the oxygen uptake except that there is dependency between O_2_ uptake and DO_2_, which is generally true only in severe anemic conditions. Generally, more pronounced changes in the brain were found for groups with lower hemoglobin (< 9.7 g/dL) [5, 45].

Both rPMRO_2_ and rCMRO_2_ raised due to RBCT, with a greater median increase in the muscle than in the brain and with a statistically significant difference between the two. This result is in disagreement with what was found in some literature related to infants where, for instance, a decrease [15] or no change (p>0.05) [39, 49] had been found in CMRO_2_ by optical methods. Likewise, no rCMRO_2_ change was found in SAH patients by PET [41] nor in children with sickle cell anemia by MRI [43]. Further research is needed to clarify these discrepancies.

As for the difference between cerebral and peripheral level response after RBCT, significant statistical difference was confirmed only for rMRO_2_ tests.

Overall, the findings for the cohort highlight an improvement in oxy- and total hemoglobin concentration and the oxygen supply, both by optical and standard methods, as expected after RBCT. Moreover, with the obtained information, it is possible to evaluate on an individual basis whether these changes pose any risks or lack them. Indeed, while anemia and RBCT are crucial for a patient’s recovery, collateral issues may arise, especially in cases of impaired cerebral autoregulation, potentially leading to stemming misery perfusion with consequent risk of cerebral ischemia. In light of the post-RBCT findings for the cohort, for instance, we did not identify any misery perfusion, as defined in Reference [28] nor a hyperperfusion or a hypoperfusion, since the rCBF did not significantly change and the rCOEF significantly decreased.

Cerebral autoregulation is often impaired in critically ill subjects [50, 51], especially in TBI [52]. In such eventuality, systemic changes may directly affect the brain and damage it [52, 53]. Therefore, it is reasonable to assume that if a robust monitor of cerebral autoregulation becomes available, it would be recommendable to utilize it frequently. In our study, the significant decrease in rCOEF along with no change in the rCBF and rCMRO_2_ suggests that the cerebral autoregulation was at play. In fact, the decrease in the rCOEF indicates that the brain was extracting less oxygen from the blood, potentially allowing for better DO_2_ to meet the metabolic demand. This is further supported by the lack of change in rCBF and rCMRO_2_, indicating a regulation mechanism to maintain consistent oxygen supply. Conversely, the significant rPBF and rPMRO_2_ increase in quadriceps along with decreased rPOEF indicate hyperemia or luxury perfusion at peripheral level. This suggests an increased supply of oxygen to the quadriceps muscle, possibly to meet increased metabolic demands, that does not imply deleterious consequences. These are rather adaptive responses to ensure adequate oxygen supply and brain protection.

The first strong limitation of this study is the small sample size. This study was a good starting point to prove the feasibility of this method and spares the need for larger clinical trials.

Once more, we recognize that our study protocol design based only on the monitoring during the first RBC bag transfusion may have been a limitation in the sense of the lower response associated to it than multiple bags. In fact, greater blood exchange may have led to potentially greater changes in our variables and could be worth further exploration to verify whether the results keep consistent over several bags or lead to significant changes.

Another important limitation is the fact that, as mentioned in the “Data evaluation” Section, the correction for the hematocrit value was taken into account in the calculation of the MRO_2_ in the analysis. However, in Reference [37] a correction method was outlined for the BFI in phantom experiments. This approach is currently under validation against MRI measurements [39] by the same group and, if confirmed *in vivo*, it would be a further step to implement in the future. Therefore, our calculations of the BFI should be considered as preliminary.

Finally, it is essential to acknowledge the potential influence of the well-known partial volume effect in our study, which is a recognized limitation when using techniques such as DCS and TRS [54, 55]. In fact, they are susceptible to variations in tissue composition within the measurement volume and the presence of different morphological tissue types and structures can lead to uncertainties in the interpretation of the acquired data, for instance the underestimation of rCBF and rOEF. While we made efforts to minimize this effect by carefully selecting measurement locations, the inherent challenge of disentangling signals from multiple tissue types, especially the extracerebral layer, was not entirely eliminated. More advanced analysis could be employed to do so. In our case, the probed tissue composition below the probes also additionally considerably varied between cerebral and peripheral measurement locations.

## Conclusion

This work focused on the compatibility of DO measurements and critically ill subjects, whose response to RBCT are more unpredictable with respect to other populations, and quantified it both at cerebral and peripheral levels. However, the applicability could be extended to other populations where anemia is a trigger to RBCT, such as during some types of surgery.

We have shown the potential usefulness of hybrid DO in RBCT management, ideally aiming at improving the RBCT decision-making in the future and paving the way to a widespread implication of hybrid DO monitors in clinical trials about RBCT. The acquired knowledge in this important clinical field could even help in personalizing RBC therapy in critical patients and enhance the quality of transfusion therapies and strategies, avoiding deleterious consequences such as hypoxia, hypoperfusion, hyperperfusion, ischemia and transfusion-associated circulatory overload.

## Data Availability

Data will be made available by the corresponding author upon reasonable request taking into account the appropriate norms for personal data privacy.

### Appendix: terminology

In this Section we clarify the distinction between the parameters derived by blood gas sample analysis from the ones by DO measurements. In fact, these two methods/fields often use the same terminology to define similar parameters although of another origin.

The oxy-hemoglobin concentration that is calculated from a blood gas sample by a co-oximeter refers to the fraction of the amount of hemoglobin present in the arterial blood that is bound to the oxygen: it corresponds to the level of oxygenated hemoglobin in blood, defining its capability to carry oxygen. In this article, we have used the acronym HgbO_2_ and Hgbr for the derived oxy- and reduced hemoglobin concentrations. As for the arterial total hemoglobin concentration, it includes the cumulative concentration of various forms of hemoglobin, such as carboxyhemoglobin and methemoglobin, in addition to the previously mentioned ones. We used Hgb to refer to it.

Oxy- and deoxy-hemoglobin concentrations that are derived from TRS measurements provide a non-invasive estimate of the content of such chromophores from deep tissue microvasculature, locally related to a specific tissue region and the underneath volume. This method does not derive these components from the blood only, rather from an average between all tissues regionally interacting with the light source and within its penetration depth. Among the usual acronyms used for optical hemoglobin concentrations, in this article we have decided to adopt the following ones in order to avoid confusion: microvascular deoxy-hemoglobin concentration ([Hhb]) and microvascular oxy-hemoglobin concentration ([HbO_2_]). The total microvascular hemoglobin concentration is calculated from the sum of these two latter and acronymized as [HbT].

We have summarized this in **Table 4**.

**Table 4:** Acronyms of the parameters related to hemoglobin concentration, to clarify which one is derived by which method.

| Acronym | Parameter | Method |
| --- | --- | --- |
| Hgb | arterial total hemoglobin concentration | blood gas analysis |
| HgbO<sub>2</sub> | arterial oxy-hemoglobin concentration | blood gas analysis |
| Hgbr | arterial reduced hemoglobin concentration | blood gas analysis |
| [HbT] | microvascular total hemoglobin concentration | diffuse topics |
| [HbO<sub>2</sub>] | microvascular oxy-hemoglobin concentration | diffuse topics |
| [Hhb] | microvascular deoxy-hemoglobin concentration | diffuse topics |

## Acknowledgements

We take this opportunity to thank the students who helped in setting up the project, Julia Scheel, Marta Grau and Lavinia Herea. We also thank Dr. J. Sahuquillo for his initial role in organizing the project. In addition, we thank all the nurses of the intensive care unit of the Vall d’Hebron hospital for allowing us to carry out the measurements in the best possible conditions, helping us with the handling of patients, extraction of blood samples and administration of transfusions. Finally, we acknowledge the useful discussions with Erin Buckley and Rowan Brothers about the hematocrit correction.

## Abbreviations

ICU: Intensive care unit
TBI: Traumatic brain injury
RBC: Red blood cells
RBCT: Red blood cells transfusion
COVID-19: Coronavirus disease 2019
HgbO_2_: Arterial oxy-hemoglobin concentration
Hgbr: Arterial reduced hemoglobin concentration
HCT: Hematocrit
TRICC: Transfusion Requirements in critical care
DO_2_: Oxygen delivery
BF: blood flow
CBF: Cerebral blood flow
PBF: Peripheral blood flow
CaO_2_: Arterial oxygen content
StO_2_: Tissue oxygen saturation
TCD: Transcranial Doppler ultrasound
OEF: Oxygen extraction fraction
COEF: Cerebral oxygen extraction fraction
POEF: Peripheral oxygen extraction fraction
[HbT]: Microvascular total hemoglobin concentration
[Hhb]: Microvascular deoxy-hemoglobin concentration
[HbO_2_ ]: Microvascular oxy-hemoglobin concentration
MRO_2_: Metabolic rate of oxygen
CMRO_2_: Cerebral metabolic rate of oxygen
PMRO_2_: Peripheral metabolic rate of oxygen
HR: heart rate
SpO_2_: Oxygen saturation
MABP: Mean arterial blood pressure
CT: Computer tomography
GCS: Glasgow coma scale
BTF: Brain trauma foundation
Hgb: Arterial total hemoglobin concentration
MCHC: Mean corpuscular hemoglobin concentration
SaO_2_: Arterial oxygen saturation
rCBF: Relative cerebral blood flow
rBF: Relative blood flow
rPBF: Relative peripheral blood flow
Δ[HbT]: Change in [HbT]
Δ[HbO_2_ ]: Change in [HbO_2_ ]
Δ[Hhb]: Change in [Hhb]
PaCO_2_: Arterial pressure of carbon dioxide
PbrO_2_: Partial pressure of tissue oxygenation
SD: Standard deviation
p: p-value
SNR: Signal-to-noise ratio
IQR: Interquartile range
ICP: Intracranial pressure
NIRS: Near-infrared spectroscopy
TRS: Time-resolved spectroscopy
DCS: Diffuse correlation spectroscopy
DO: Diffuse optics rOEF: Relative oxygen extraction fraction
rCOEF: Relative cerebral oxygen extraction fraction
rPOEF: Relative peripheral oxygen extraction fraction
rMRO_2_: Relative metabolic rate of oxygen
rCMRO_2_: Relative cerebral metabolic rate of oxygen
rPMRO_2_: Relative peripheral metabolic rate of oxygen
PaO_2_: Partial pressure of oxygen
SaO_2_: Arterial oxygen saturation
MRI: magnetic resonance imaging
PET: positron emission tomography
SAH: subarachnoid hemorrhage
CBV: cerebral blood volume
NA: not available

## Ethics approval and consent to participate

The study obtained clearance by the ethical committee of Vall D’Hebron University Hospital (PR(AG)160/2017) and was conducted following the Declaration of Helsinki [56]. Signed informed consent was obtained before the measurements either by the patient or a legal representative.

## Authors’ contributions

S.T. had full access to all data in the study and took responsibility for the integrity of the data and the accuracy of the analysis. S.T. performed all optical measurements, data analysis and wrote the manuscript. S.T., M.K. and J.B.F. built the optical device used for the acquisitions. L.E. helped during the data acquisition. A.R., A.F.J. and S.V.A. were responsible of the patient recruitment. F.M. provided clinical information about the patients. M.A.P. and T.D. were responsible of the study design, contributed to the results interpretation and writing of the manuscript. All authors read and approved the final manuscript.

## Footnotes

[1] Details about the terminology and acronyms used in this article are reported in the **Appendix**.

